# Large Language Model Influence on Management Reasoning: A Randomized Controlled Trial

**DOI:** 10.1101/2024.08.05.24311485

**Authors:** Ethan Goh, Robert Gallo, Eric Strong, Yingjie Weng, Hannah Kerman, Jason Freed, Joséphine A. Cool, Zahir Kanjee, Kathleen P. Lane, Andrew S. Parsons, Neera Ahuja, Eric Horvitz, Daniel Yang, Arnold Milstein, Andrew P.J Olson, Jason Hom, Jonathan H Chen, Adam Rodman

**Author notes:** These authors contributed equally to this work (Ethan Goh, Robert Gallo). These authors contributed equally to this work (Jason Hom, Jonathan H Chen, Adam Rodman). Corresponding Author: Jonathan H Chen, MD, PhD, Assistant Professor, Center for Biomedical Informatics Research, Stanford Department of Medicine, 500 Pasteur Dr Stanford, CA 94305.

## Abstract

**Importance:** Large language model (LLM) artificial intelligence (AI) systems have shown promise in diagnostic reasoning, but their utility in management reasoning with no clear right answers is unknown.

**Objective:** To determine whether LLM assistance improves physician performance on open-ended management reasoning tasks compared to conventional resources.

**Design:** Prospective, randomized controlled trial conducted from 30 November 2023 to 21 April 2024.

**Setting:** Multi-institutional study from Stanford University, Beth Israel Deaconess Medical Center, and the University of Virginia involving physicians from across the United States.

**Participants:** 92 practicing attending physicians and residents with training in internal medicine, family medicine, or emergency medicine.

**Intervention:** Five expert-developed clinical case vignettes were presented with multiple open-ended management questions and scoring rubrics created through a Delphi process. Physicians were randomized to use either GPT-4 via ChatGPT Plus in addition to conventional resources (e.g., UpToDate, Google), or conventional resources alone.

**Main Outcomes and Measures:** The primary outcome was difference in total score between groups on expert-developed scoring rubrics. Secondary outcomes included domain-specific scores and time spent per case.

**Results:** Physicians using the LLM scored higher compared to those using conventional resources (mean difference 6.5 %, 95% CI 2.7-10.2, p<0.001). Significant improvements were seen in management decisions (6.1%, 95% CI 2.5-9.7, p=0.001), diagnostic decisions (12.1%, 95% CI 3.1-21.0, p=0.009), and case-specific (6.2%, 95% CI 2.4-9.9, p=0.002) domains. GPT-4 users spent more time per case (mean difference 119.3 seconds, 95% CI 17.4-221.2, p=0.02). There was no significant difference between GPT-4-augmented physicians and GPT-4 alone (-0.9%, 95% CI -9.0 to 7.2, p=0.8).

**Conclusions and Relevance:** LLM assistance improved physician management reasoning compared to conventional resources, with particular gains in contextual and patient-specific decision-making. These findings indicate that LLMs can augment management decision-making in complex cases.

**Trial Registration:** ClinicalTrials.gov Identifier: NCT06208423;

https://classic.clinicaltrials.gov/ct2/show/NCT06208423

**Key Points:** *Question:* Does large language model (LLM) assistance improve physician performance on complex management reasoning tasks compared to conventional resources?

*Findings:* In this randomized controlled trial of 92 physicians, participants using GPT-4 achieved higher scores on management reasoning compared to those using conventional resources (e.g., UpToDate).

*Meaning:* LLM assistance enhances physician management reasoning performance in complex cases with no clear right answers.

## Introduction

Large language models (LLMs) show considerable abilities in diagnostic reasoning, outperforming previous artificial intelligence (AI) models as well as human physicians in their ability to construct helpful differential diagnoses, explain reasoning, and collect historical information from standardized patients.^1–5^ LLMs have not yet shown to perform similarly in management reasoning, which encompasses decision making around treatment, testing, patient preferences, social determinants of health, and cost-conscious care, all while managing risk.^6–8^

While there is overlap, clinical reasoning is often considered to comprise both diagnostic and management reasoning. The study of diagnostic reasoning has a century-long history with numerous metacognitive frameworks and assessment methods, while management reasoning processes represent a comparatively recent area of study.^9–11^ Current frameworks in management reasoning include context-dependent concepts such as shared decision making, dynamic relationships and competing priorities between medical systems and individuals, the physician-patient relationship, and time constraints inherent in modern clinical encounters.^8,12^ In contrast to diagnostic reasoning, which can be thought of as a classification task with often a single right answer, management reasoning often has no right answers and involves weighing trade-offs between inherently risky courses of action; even inaction through “watchful waiting” is a deliberate choice with potential risks and benefits. From a cognitive psychology perspective, management reasoning often utilizes heuristics called management scripts which allow clinicians to quickly make decisions.^13^ However, these scripts are susceptible to the same fallibilities that affect other domains of human reasoning. With few exceptions, these scripts must be adapted to specific situations to balance all factors that influence management reasoning, as well as continually updated with new and emerging information.

Previous generations of non-LLM AI systems can improve human management decisions in some situations, especially when a human user treats an AI suggestion as a second opinion.^14^ One of the theoretical strengths of LLMs is their ability to serve as a cooperation partner, augmenting human cognition.^15^ LLMs may offer differing points of view that would assist in aligning patients’ and clinicians’ values and goals into a cohesive plan, but this has not been experimentally tested. We designed a prospective randomized-controlled trial to assess whether physicians using an LLM performed better than physicians using standard resources on a series of complex clinical management questions. We then compared physician answers to the output of the LLM (without a human) alone.

## Methods

### Participants

We recruited practicing attending physicians and resident physicians with training in a general medical specialty (internal medicine, family medicine, or emergency medicine) from Stanford University, Beth Israel Deaconess Medical Center, the University of Virginia via e-mail. Informed consent was obtained prior to enrollment and randomization. This study was reviewed and determined to be exempt by institutional review boards at Stanford University, Beth Israel Deaconess Medical Center, and the University of Virginia. Small groups of participants were proctored by study coordinators either remotely or at an in-person computer laboratory. Sessions lasted for one hour. Resident physicians were offered $100 and attending physicians were offered $200 for completing the study.

### Clinical case vignette construction

We constructed our cases from the series of *Grey Matters* from the American College of Physicians podcast “Core IM.”^16^ As these cases were adapted specifically for this study, they were not available to either GPT-4 or the participants prior to our study. Each of these cases was constructed via a panel of subspecialty and generalist experts (including authors AR, ZK, ES, JH, and AP) to explore how physicians make decisions when there are no clear right answers. We intentionally chose a selection of cases that would explore the breadth of general medicine management decision making. Through initial pilot studies (not included in the analysis), we determined that no participant finished more than 5 cases within one hour, in line with standardized tests of physician reasoning such as licensing exams and observed structured clinical examinations.^17,18^

### Development of scoring rubrics

The paramount challenge in the evaluation of management reasoning is the relatively wide variety of reasonable answers depending on contextual factors.^19,20^ Unlike a confirmed pathological final diagnosis, there is often a range of acceptable answers for management reasoning. To capture this nuance of a variety of management perspectives, for each case, we convened an expert group of five individuals – a member of the study team, two generalists, and two subspecialists in the field applicable to the case. Through an iterative modified Delphi process, we refined management rubrics to score each case.^21^ These rubrics were designed to be as thorough as possible for the specific case, while also acknowledging that considerable variation of acceptable management was possible (eSupplement for example of case and rubric). Because of this, scores on the rubrics do not comport with standard cut offs from educational interventions (e.g., 40% neither reflects a “passing” or “failing” score, only a percentage of the total possible points possible in a comprehensive rubric; points are given for all answers determined reasonable by the panel and while a higher score reflects a more comprehensive answer, there is no clear cut off for high quality care). Each of these rubrics were tested in two pilot groups and further refined with user feedback. Because there is not often a clear divide between the diagnostic and management domains of clinical reasoning, each question was independently labeled by two members of the study team (EG and HK) as reflecting case-specific reasoning or more generalized clinical reasoning that did not require case-specific information. Case questions were similarly categorized as representing a diagnostic decision (for example, a differential for an incidentally-found lung nodule), a management decision (for example, the contextual factors that drive next steps in the work up of a lung nodule) or knowledge recall (the risk factors that make a lung nodule more likely to be malignancy). There was complete agreement on these labels.

### Study design

We employed a prospective, randomized, single-blind (to the rater) study design with participants randomized to either use GPT-4 via the ChatGPT Plus [OpenAI, San Francisco, CA] interface or the conventional resources group (Figure 1). To mirror real-world implementation, participants received GPT-4 training comparable to current live deployments in clinical settings.^22^ This included basic instruction on system access and use, as well as live technical support throughout the study from a proctor.

**Figure 1:**
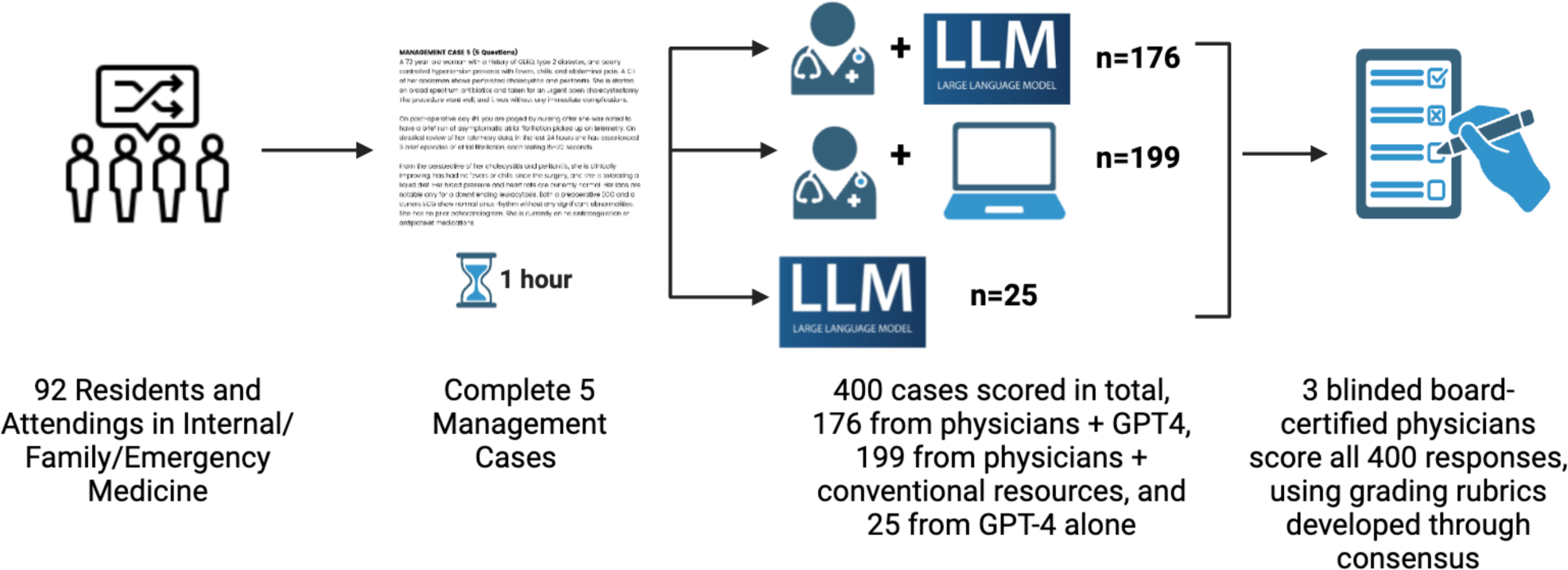
Study Flow Diagram. *92 practicing attending physicians and residents with training in internal medicine, family medicine, or emergency medicine. Five expert-developed cases were presented, with scoring rubrics created through a Delphi process. Physicians were randomized to use either GPT-4 via ChatGPT Plus in addition to conventional resources (e.g., UpToDate, Google), or conventional resources alone. The primary outcome was difference in total score between groups on expert-developed scoring rubrics. Secondary outcomes included domain-specific scores and time spent per case*.

Both groups were instructed that they could use any point-of-care resources they normally use in clinical practice, such as UpToDate [Wolters Kluwer, Philadelphia, PA], Epocrates [Athenahealth, Watertown, MA], and other internet resources. The control group was instructed not to use any LLMs (e.g., ChatGPT, Claude, Bard/Gemini). We instructed the participants to finish as many of the five cases as they could in an hour, prioritizing quality of responses over completing all cases. The study was conducted using a Qualtrics (Qualtrics, Seattle, WA) survey tool; participants received the cases in sections before moving on. They were not able to change their answers to prior prompts as new pieces of information were introduced.

### Prompt design for LLM-only arm

For the LLM-only arm, we used established principles of prompt design to iteratively develop a zero-shot prompt by copy-pasting the management cases along with questions (Appendix).^23^ Each prompt was run five times, and the results from the five runs were included for blinded grading alongside the human outputs before any unblinding or data analysis.

### Rubric validation

Two preliminary sets of data from 10 individuals total were collected for validation of the rubrics. The three graders (AR, ES, and KL) independently graded these two data sets. They then met in person and came to consensus on grading these two validation sets. After data collection was complete, each case was graded independently by two of three graders who were blinded to group assignment. When scorers disagreed (predefined as a difference of >10% of the final score), they met to discuss differences in their assessments and to seek consensus. Final diagnosis scoring was adjudicated by two reviewers to obtain agreement for the secondary outcome of diagnostic accuracy. We calculated a weighted Cohen’s kappa to show concordance in grading, both for each individual case and for all cases pooled together.

### Study outcomes

The primary study outcome was the mean score for each of the groups. Secondary outcomes included the scores in predefined domains of the rubrics, including management, knowledge recall and diagnostic domains, case specificity or generality of decisions, as well as time spent on cases.

### Statistical methods

The target minimum sample size of 84 participants was pre-specified based on a power analysis using preliminary data of 13 cases among three participants, scored prior to study enrollment, corresponding to an expected 252 to 336 cases completed (3–4 cases per participant). This minimum target sample size ensured sufficient power (> 80%) for both the primary outcome and time spent on cases as the secondary outcome. All analyses were at the case level, clustered by the participant. In the primary analysis, we only included cases with completed responses – that is, answered up to the final question. Generalized mixed-effect models were applied to assess the difference in the primary and secondary outcomes of the GPT-4 group compared to the conventional resources only group. A random-effect for the participant was included in the model to account for the potential correlation between cases for a participant. Additionally, a random effect for cases was included to account for any potential variability in difficulty across cases. All statistical analysis was performed using R v4.3.2 (R Foundation for Statistical Computing, Vienna, Austria). Statistical significance was based on a p value <0.05.

## Results

We enrolled 92 physicians to participate in the study conducted from 30 November 2023 to 21 April 2024. Participants were randomized evenly between the LLM group and conventional resources group (Table 1). 73% (67/92) were attendings; 27% (25/92) were residents. 74% (68/92) specialized in internal medicine, 20% (18/92) emergency medicine, and 6.5% (6/92) family medicine. The mean time in practice of all physicians was 7.6 years; median 5.8 years (IQR: 3.0 to 9.0 years). Only 24% (22/92) self-described themselves as frequent users of LLMs; 20.8% (19/92) had either only used it once or never used it.

**Table 1:** Participant Characteristics by Randomized Groups.

| Variable | Overall,<br>N = 92 | Physicians+LLM,<br>N = 46 | Physicians +<br>Conventional<br>Resources only,<br>N = 46 | SMD |
| --- | --- | --- | --- | --- |
| Career Stage |  |  |  | 0.05 |
| Attending | 67 (73%) | 34 (74%) | 33 (72%) |  |
| Resident | 25 (27%) | 12 (26%) | 13 (28%) |  |
| Specialty |  |  |  | 0.22 |

|  |  |  | Physicians +<br>Conventional<br>Resources only, |  |
| --- | --- | --- | --- | --- |
| Variable | Overall,<br>N = 92 | Physicians+LLM,<br>N = 46 | N = 46 | SMD |
| Internal Medicine | 68 (74%) | 36 (78%) | 32 (70%) |  |
| Emergency Medicine | 18 (20%) | 8 (17%) | 10 (22%) |  |
| Family Medicine | 6 (6.5%) | 2 (4.3%) | 4 (8.7%) |  |
| **Years in Medical**<br>**Training** |  |  |  | -0.02 |
| Mean (SD) | 7.6 (7.1) | 7.6 (7.9) | 7.7 (6.3) |  |
| Median [IQR] | 5.8<br>[3.0 to 9.0] | 5.0<br>[3.1 to 8.8] | 6.0<br>[3.0 to 9.8] |  |
| **Past GPT Experience** |  |  |  | 0.11 |
| I use it frequently<br>(weekly or more) | 22 (24%) | 11 (24%) | 11 (24%) |  |
| I use it occasionally<br>(more than once per<br>month but less than<br>weekly) | 28 (30%) | 15 (33%) | 13 (28%) |  |
| Variable | Overall,<br>N = 92 | Physicians+LLM,<br>N = 46 | N = 46 | SMD |
| I use it rarely (less than<br>once per month) | 23 (25%) | 11 (24%) | 12 (26%) |  |
| I've used it once ever | 9 (9.8%) | 4 (8.7%) | 5 (11%) |  |
| I've never used it before | 10 (11%) | 5 (11%) | 5 (11%) |  |

**Table 2:** Comparisons of the Primary and Secondary Outcomes by Physicians with LLM and with Conventional Resources Only (Scores standardized to 0-100)

|  |  | Physicians +<br>Conventional<br>Resources Only, | Difference between<br>Physicians+GPT-4 and<br>Physicians+Conventional<br>Resources Only<br>(based on Generalized Mixed Effect<br>Model, 95% Confidence Interval, *p*-<br>*value*) |
| --- | --- | --- | --- |
| Outcomes | Physicians+LLM,<br>N = 178<sup>*†*</sup> | Resources Only,<br>N = 197<sup>*†*</sup> |  |
| Primary Outcome |  |  |  |
| Total Score (n) | 178 | 197 | 6.5 (2.7 to 10.2), *p*<0.001 |
| Mean (SD) | 43.0 (17.3) | 35.7 (15.5) |  |
| Median [IQR] | 41.3<br>[30.6 to 54.1] | 34.4<br>[22.5 to 47.8] |  |
| Secondary Outcomes |  |  |  |
| Management (n) | 178 | 197 | 6.1 (2.5 to 9.7), *p*=0.001 |
| Mean (SD) | 40.5 (19.1) | 33.4 (17.3) |  |
| Median [IQR] | 37.5<br>[26.8 to 52.4] | 30.0<br>[19.3 to 45.5] |  |
| **Factual (n)** | 69 | 78 | 9.6 (-3.1 to 22.3), p=0.14 |
| Mean (SD) | 62.9 (37.6) | 53.8 (39.6) |  |
| Median [IQR] | 75.0<br>[37.5 to 100.0] | 56.2<br>[15.6 to 100.0] |  |
| **Diagnostic (n)** | 72 | 77 | 12.1 (3.1 to 21.0), p=0.009 |
| Mean (SD) | 56.8 (37.6) | 45.8 (26.7) |  |
| Median [IQR] | 66.7<br>[29.2 to 83.3] | 50.0<br>[33.3 to 66.7] |  |
| **Specific (n)** | 178 | 197 | 6.2 (2.4 to 9.9), p=0.002 |
| Mean (SD) | 42.4 (20.2) | 34.9 (17.9) |  |
| Median [IQR] | 42.6<br>[28.1 to 57.4] | 35.2<br>[20.8 to 48.5] |  |
| **General (n)** | 70 | 80 | 3.3 (-1.3 to 7.9), p=0.2 |
| Mean (SD) | 29.4 (15.0) | 26.5 (13.0) |  |
| Median [IQR] | 27.3<br>[18.2 to 39.8] | 24.6<br>[17.5 to 33.3] |  |
| **Time Spent in Seconds (n)** | 178 | 197 | 119.3 (17.4 to 221.2), p=0.022 |
| Mean (SD) | 801.5 (417.2) | 690.2 (372.4) |  |
| Median [IQR] | 719.8<br>[514.6 to 1,010.2] | 570.9<br>[452.9 to 814.9] |  |

From these 92 physicians, 400 cases were scored in total, 176 from the group of physicians using the LLM, 199 from physicians using only conventional resources, and 25 from the LLM alone. Three graders agreed on the scoring of 328 of 400 cases (82%), with a pooled κ of 0.80, reflecting substantial agreement (case 1 κ = 0.58, case 2 κ = 0.83, case 3 κ = 0.82, case 4 κ = 0.90, case 5 κ = 0.89).

### Management performance

Physicians randomized to use the LLM performed better than the control group (43.0% compared to 35.7%, difference: 6.5%, 95% CI: 2.7% to 10.2%, p<0.001) (Figure 2). The LLM by itself scored comparably to humans using the LLM (43.7% versus 43.0%, difference: 0.9%, 95% CI: -7.2% to 9.0%, p =0.80), while trending towards scoring higher than humans using conventional resources (43.7% versus 35.7%, difference: 7.3%, 95% CI: -0.7% to 15.4%, p =0.074).

**Figure 2.**
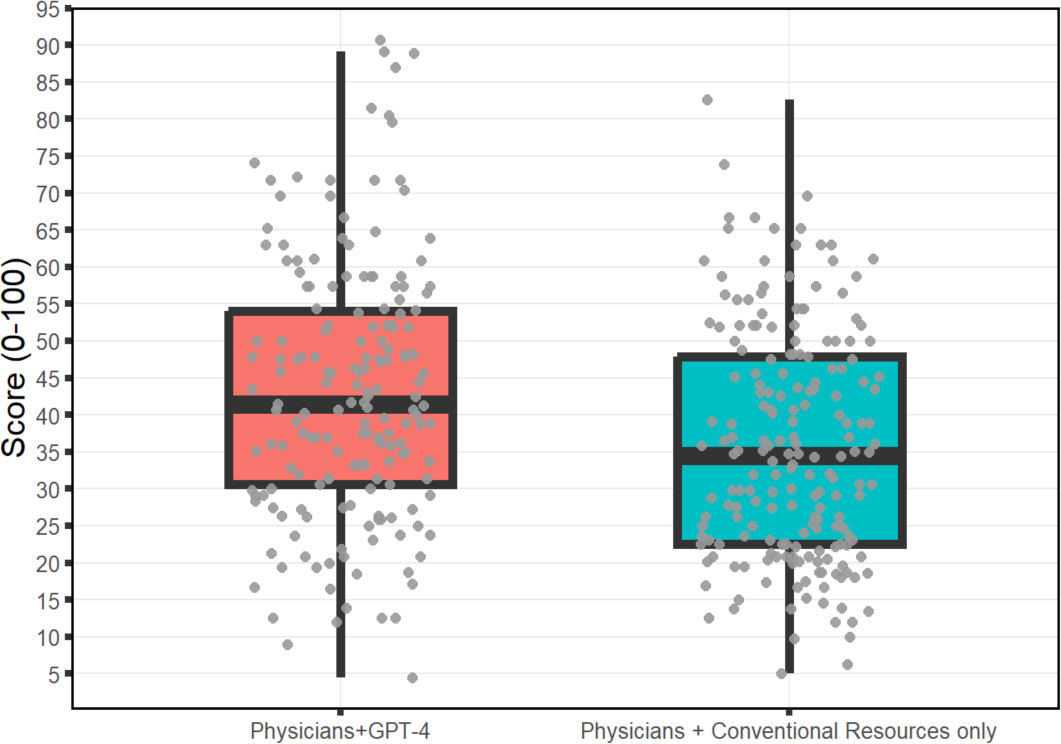
**Comparisons of the Primary Outcome by Physicians with LLM and with Conventional Resources Only (Total score standardized to 0-100)**

### Question domain subgroups

The physicians using LLM scored better than conventional resources alone in questions explicitly testing management decisions (40.5% versus 33.4%, difference: 6.1%, 95% CI: 2.5% to 9.7%, p=0.001), questions testing diagnostic decisions (56.8% versus 45.8%, difference: 12.1%, 95% CI: 3.1% to 21%, p = 0.009), and context-specific questions (42.4% versus 34.9%, difference: 6.2%, 95% CI: 2.4% to 9.9%, p = 0.002). While we were unable to detect a difference between the two groups in factual recall (62.9% versus 53.8%, difference: 9.6%, 95% CI: -3.1% to 22.3%, p = 0.14) and general management knowledge (29.4% versus 26.5%, difference: 3.3, 95% CI: -1.3% to 7.9%, p = 0.2), they were overall directionally similar to the other subdomains.

### Time

Physicians randomized to use GPT-4 spent 111.3 seconds more on each case (801.5 seconds versus 690.2, difference: 119.3 seconds, 95% CI: 17.4 to 221.2, p = 0.022).

## Discussion

In this randomized controlled trial, the availability of an LLM improved physician management reasoning compared to conventional resources only, with comparable scores between physicians randomized to use AI and the AI alone. This suggests a future use for LLMs as a helpful adjunct to clinician judgment. From a cognitive psychology perspective, it is surprising that an LLM would enhance management reasoning. The abilities of LLMs to make diagnostic decisions likely results from their underlying token prediction architecture and its similarities to how physicians cluster and activate semantic illness scripts in making diagnoses.^24^ Management scripts, on the other hand, are highly contextual and individualized, and include many factors outside of the biomedical encounter. Thus, the best decision for a patient in a given situation may be different than another patient with the exact same condition in a different context. For example, the appropriate management of an incidentally discovered 2.0 centimeter upper lobe lung nodule in a hospitalized inpatient might be immediate biopsy in a patient unlikely to follow up; scheduled outpatient biopsy in a health system capable of organizing and ensuring continuity; outpatient PET scan in a patient reticent to undergo an invasive procedure; or serial imaging in a patient with limited life expectancy. The knowledge that such a large nodule in the upper lobe has a high chance of representing malignancy is only the first step in formulating a follow up plan – patient preferences, knowledge of the healthcare system, and the patient’s social situation are equally important factors.

The group using the LLM also spent more time solving cases, another surprising finding, and in contrast to findings in diagnostic reasoning.^4,5^ Interacting with the LLM may have served as a “time out” to drive better decisions and consider the patient’s context. For example, physicians using the LLM often considered patient and other contextual factors in management decisions, as well as exhibiting apparent empathy to other providers and patients in difficult situations. We suspect that some of these emergent abilities come from the fine-tuning process called reinforcement learning through human feedback (RLHF), in which empathetic and patient-centered responses are rated as favorable by humans.^25^ Similar to studies showing increased empathetic communication phrasing from LLMs to patient queries, this study indicates that LLMs may influence physicians to better consider human factors in their management reasoning.^26–28^ Improved humanistic and patient-centered behaviors of clinicians when they collaborate with an LLM is an important and even reassuring finding even at the expense of taking more time.

This study has multiple limitations. First, all our cases are clinical vignettes, based on, but not actual patient cases. While our scoring rubrics show substantial inter-rater reliability, validity evidence for this rubric has not been gathered outside this study. Only potentially correct answers were given credit, while wrong answers were not penalized. LLM-based outputs are often significantly longer than human responses, and it is thus possible that physicians in the AI group scored better merely through verbosity alone.

With only five cases expected for participants to complete within a one-hour session, we intentionally selected content to represent a breadth of general medicine, in line with standardized evaluations such as objective structured clinical examinations. A wider variety of cases could show different outcomes. Finally, we provided only basic training on use of LLMs to either group as well as technical support. While evidence suggests prompting strategies can dramatically improve model performance on medical tasks, we intentionally chose to mimic current strategies around LLM deployment in healthcare settings, which have been provided with minimal formal training on prompting strategies.^22,29^

This study found that the addition of LLM AI assistance improves physician management reasoning compared to conventional resources. Early implementations of LLMs into healthcare have largely been directed at clerical clinical workflows, including portal messaging and ambient listening. Decision support – even in something as complex as management reasoning – appears to be another domain requiring further research and development to ensure safe and effective integration of such tools for enhanced patient care.

## Data Availability

Example case vignettes, questions, and grading are included in the supplement. GPT-4 transcript chat logs, raw score table, and individual survey responses are available upon request.

## Contributions

Ethan Goh (co-first author) - Study design, data acquisition, data interpretation, manuscript preparation, critical revision

Robert Gallo (co-first author) - Study design, data acquisition, data interpretation, manuscript preparation, critical revision

Eric Strong - Study design, data acquisition, data interpretation

Yingjie Weng - Study design, data interpretation

Hannah Kerman - Study design, data acquisition, data interpretation

Jason Freed - Study design, data interpretation

Josephine Cool - Study design, data interpretation

Zahir Kanjee - Study design, data interpretation

Kathleen Lane - Study design, data interpretation

Andrew Parsons - Study design, data interpretation

Daniel Yang - Study design, data interpretation

Arnold Milstein - Funding and administrative support

Neera Ahuja - Funding and administrative support

Eric Horvitz - Study design, critical revision

Andrew Olson - Study design, data interpretation

Jason Hom - Study design, data analysis, data interpretation, critical revision, supervision, funding and administrative support

Jonathan Chen (co-last author) - Study design, data analysis, data interpretation, critical revision, supervision, funding and administrative support

Adam Rodman (co-last author) - Study design, data analysis, data interpretation, critical revision, supervision

