## Supplement 1 to 7 for "Large Language Model Influence on Management Reasoning: A Randomized Controlled Trial"

**SUPPLEMENTARY MATERIALS**

**Supplement 1: Study Flow Diagram**

**Supplement 2: Example of a Management Case and Rubric**

**Supplement 3: Example of High-scoring vs. Low-scoring Responses**

**Supplement 4: GPT Prompt and Responses for Example Management Case**

**Supplement 5: Comparisons of the Primary and Secondary Outcomes by GPT Alone vs Physicians with GPT-4 and Physicians with Conventional Resources Only (Scores standardized to 0-100)**

**Supplement 6: Comparisons of the Primary Outcome by GPT Alone vs Physicians with GPT-4 and with Conventional Resources Only (Total score standardized to 0-100)**

**Supplement 7: Comparisons of Time Spent by Physicians with GPT-4 and Physicians with Conventional Resources Only**

### Supplement 1: Study Flow Diagram

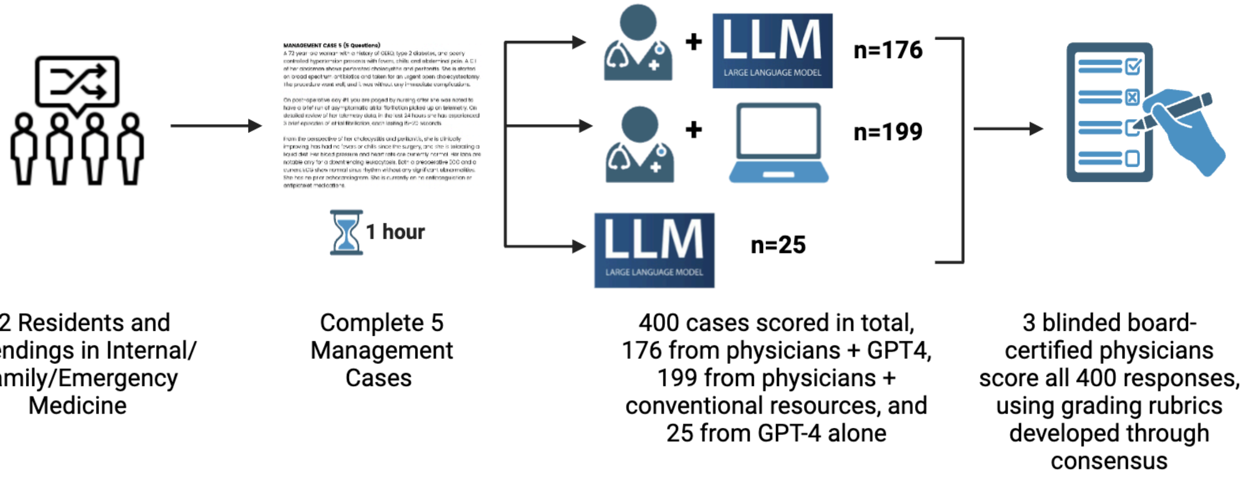

*92 practicing attending physicians and residents with training in internal medicine, family medicine, or emergency medicine. Five expert-developed cases were presented, with scoring rubrics created through a Delphi process. Physicians were randomized to use either GPT-4 via ChatGPT Plus in addition to conventional resources (e.g., UpToDate, Google), or conventional resources alone. The primary outcome was difference in total score between groups on expert-developed scoring rubrics. Secondary outcomes included domain-specific scores and time spent per case.*

### Supplement 2: Example of a Management Case and Rubric

**Case #4**
**23 total points possible**

**CLINICAL VIGNETTE (PART 1):**

A 72 year-old woman with a history of GERD, type 2 diabetes, and poorly controlled hypertension presents with fevers, chills, and abdominal pain. A CT of her abdomen shows perforated cholecystitis and peritonitis. She is started on broad spectrum antibiotics and taken for an urgent open cholecystectomy. The procedure went well, and it was without any immediate complications.

On post-operative day #1, you are paged by nursing after she was noted to have a brief run of asymptomatic atrial fibrillation picked up on telemetry. On detailed review of her telemetry data, in the last 24 hours she has experienced 3 brief episodes of atrial fibrillation, each lasting 15-20 seconds.

From the perspective of her cholecystitis and peritonitis, she is clinically improving, has had no fevers or chills since the surgery, and she is tolerating a liquid diet. Her blood pressure and heart rate are currently normal. Her labs are notable only for a downtrending leukocytosis. Both a preoperative ECG and a current ECG show normal sinus rhythm without any significant abnormalities. She has no prior echocardiogram. She is currently on no anticoagulation or antiplatelet medications.

**QUESTION 1:**

In addition to continuing telemetry monitoring, would you recommend any additional monitoring, testing, or treatment at this time? If so, what?

**This question has a total of <u>4 points</u>.**

| | |
|---|---|
| Prophylactic anticoagulation to prevent VTE/DVT (e.g. enoxaparin 40mg daily) – Credit for either correct labeling of the intervention, or correct med + dose combination | 1 point |
| TSH | 1 point |
| Inclusion of up to 2 of the following:<br>● Echocardiogram<br>● Electrolyte assessment and/or repletion<br>● Evaluation for additional triggers (e.g. pain, constipation, etc…) | 2 points max (1 point each) |

**CLINICAL VIGNETTE (PART 2):**

There are no episodes of atrial fibrillation on either post-operative days #2 or #3. However, on post-operative day #4, the day prior to anticipated discharge, the patient again develops another episode on telemetry that is confirmed by ECG. This episode is also completely asymptomatic. It lasts two hours, during which the ventricular rate ranges from 70 to 105 beats/min. Her blood pressure is not affected. It spontaneously converts back to normal sinus rhythm.

**QUESTION 2:**

Would you recommend to this patient that she be started on therapeutic anticoagulation?
What factors would influence your decision?

**This question has a total of <u>9 points</u>.**

Decision on anticoagulation

| | |
|---|---|
| Yes, start therapeutic anticoagulation | 2 points |

Factors which influence the decision

| | |
|---|---|
| Patient preference and/or risk tolerance | 2 points |
| ● CHA2DS2-VASc or CHADS2 score<br>● Listing only 1-2 component of these scoring systems separately, without mentioning the score itself | 2 points<br>1 point |
| Inclusion of up to 3 of the following 5 factors:<br>● HAS-BLED score<br>● Duration of a-fib episodes<br>● # of post-op episodes<br>● Information potentially obtained from an echo | 3 points max (1 point for each) |

| |
|---|
| • A-fib burden (i.e. % of time spent in a-fib) |

**QUESTION 3:**

If the decision was made to start the patient on therapeutic anticoagulation on hospital day #4 (i.e. immediately after the asymptomatic, 2 hour long episode of a-fib), what would be the best choice for anticoagulation?

**This question has a total of <u>3 points</u>.**

| | |
|---|---|
| Apixaban, rivaroxaban, edoxaban, dabigatran, therapeutic enoxaparin | 3 points |
| Unfractionated heparin gtt | 1 point |

**QUESTION 4:**

**This question has a total of <u>7 points</u>.**

The patient is about to be discharged. Would you discharge her with an ambulatory ECG monitor?

| | |
|---|---|
| Yes | 1 point |

If so, what type of monitor would you choose?

| | |
|---|---|
| Holter and/or Ziopatch (i.e. “14 day Holter”) | 1 point |

What factors should be considered in the choice of monitor for this patient?

| | |
|---|---|
| Inclusion of up to 2 of the following:<br>• Cost of monitor<br>• Ease of insurance approval<br>• Duration of monitoring period<br>• Likelihood of the patient experiencing a lethal arrhythmia | 2 points max (1 point each) |

What would you do with the information from the monitor?

| | |
|---|---|
| Determine anticoagulation plan (i.e. whether or not the patient requires anticoagulation, or the duration of anticoagulation) | 1 point |
| Inclusion of up to 2 of the following:<br>• Determine if the patient requires rate control<br>• Determine if the patient requires rhythm control | 2 points max (1 point each) |

| |
|---|
| • Determine if the patient requires more diagnostic testing |

**Supplement 3: Example of High-scoring vs. Low-scoring Responses**

***Example of High-scoring response***

**Question 1. In addition to continuing telemetry monitoring, would you recommend any additional monitoring, testing, or treatment at this time? If so, what?**

I'd check her electrolytes and make sure hypoK or hypomag weren't contributing to her tacchyarrythmia. I'd also get trops and an NT-proBNP to ensure there wasn't an inciting cardiac event triggering her afib, and TSH to ensure she wasn't hyperthyroid. I'd consider an echo. I'd ensure she was placed on tele as well. It's hard to say whether she is having paroxysmal afib in the setting of her acute illness or whether it has been happening at home before she was admitted. I would therefore discharge her with a ziopatch and cardiology follow-up to help make that distinction. She also has a CHA2DS2-VASc score of 4, which puts her at moderate-high risk of cardioembolic stroke and she thus warrants anticoagulation. I'd talk to her and see if she has any bleeding risk factors - if not, I'd likely start her on apixaban. I'd also provide situational awareness to the rest of her inpatient team that she could be started on metoprolol or amiodarone if she were to recur into afib but with RVR.

**Question 2. Would you recommend to this patient that she be started on therapeutic anticoagulation?**

Yes - she's at moderate-high risk of cardioembolic stroke. This could always be peeled off in the outpatient setting if she undergoes additional cardiac monitoring with no demonstrated afib in case her afib here is triggered by stress from recent surgery, but regardless of how permanent her afib is she is a high stroke risk and I would want to minimize this.

**Question 2b. What factors would influence your decision?**

Her willingness to take an anticoagulant, her ability to pay for a DOAC, her willingness to accept the hassles of being on warfarin or lovenox if she can't pay for a DOAC, her bleeding risk (does she fall, does she have any recent bleeds - could use HAS-BLED to better quantify this), her hemoglobin, whether she's had a stroke recently, whether I expect her surgical site to bleed, whether she can follow up with a physician after this hospitalization, whether anticoagulation would have a harmful interaction with any of her current medications.

**Question 3. If the decision was made to start the patient on therapeutic anticoagulation on hospital day #4 (i.e. immediately after the asymptomatic, 2 hour long episode of a-fib), what would be the best choice for anticoagulation?**

Apixaban

**Question 4a. The patient is about to be discharged. Would you discharge her with an ambulatory ECG monitor?**

Yes - I would want to monitor whether her afib is only present in the provoked post-op setting vs. if she has it recurrently at home, even when her body is no longer experiencing acute illness.

**Question 4b. If so, what type of monitor would you choose?**

Ziopatch

**Question 4c. What factors should be considered in the choice of monitor for this patient?**

Affordability and insurance coverage, accuracy of the monitor, whether the cardiologist she sees following hospitalization has the IT systems in place to receive the readout of the monitor, her ability to incorporate the monitor into her daily life and adhere to wearing it, the duration of monitoring that is most clinically appropriate (I'd go for 30 days).

**Question 4d. What would you do with the information from the monitor?**

Confirm whether she is having ongoing episodes of afib and if so better characterize its frequency, duration, and pattern; correlate any symptoms with her afib; guide decision-making around anticoagulation; selection/initiation of rhythm or rate control agents; and consideration for more permanent treatment options like cardiac ablation or Watchman device placement in case she wanted to get off of anticoagulation.

**Total score: 21 (of 23)**

***Example of a low-scoring response***

**Question 1. In addition to continuing telemetry monitoring, would you recommend any additional monitoring, testing, or treatment at this time? If so, what?**

TTE and trop/CKMB. Can also consider a heart monitor such as a ziopatch upon d/c.

**Question 2. Would you recommend to this patient that she be started on therapeutic anticoagulation?**

Given recent surgery and current duration of episodes, at this time I would not, however I would place a monitoring device and insure soon outpt f/u w/ cardiology

**Question 2b. What factors would influence your decision?**

Medical hx, activity level, d/c labs, close f/u.

**Question 3. If the decision was made to start the patient on therapeutic anticoagulation on hospital day #4 (i.e. immediately after the asymptomatic, 2 hour long episode of a-fib), what would be the best choice for anticoagulation?**

Apixaban or Xarelto

**Question 4a. The patient is about to be discharged. Would you discharge her with an ambulatory ECG monitor?**

Yes

**Question 4b. If so, what type of monitor would you choose?**

I would consider a ziopatch or monitor such as that one.

**Question 4c. What factors should be considered in the choice of monitor for this patient?**

Risk factors, medical conditions, overall activity level.

**Question 4d. What would you do with the information from the monitor?**

Have it read by cardiology during pt f/u appt. It can also help evaluate for eventual need of AC or rhythm control medication

**Total score: 8 (of 23)**

**<u>Supplement 4: GPT Prompt and Responses for Example Case</u>**

| GPT-4 Prompt | GPT-4 Responses |
|---|---|
| You are an experienced internal medicine physician undergoing a test of your medical management skills. I am going to give you a case with multiple questions. These are not real patients but are examples purely to test your management reasoning. Please answer these questions as best as possible. If the question is a yes or no question, please answer it directly. Please number your responses the same way they are numbered in the question stem.<br><br>A 72 year-old woman with a history of GERD, type 2 diabetes, and poorly controlled hypertension presents with fevers, chills, and abdominal pain. A CT of her abdomen shows perforated cholecystitis and peritonitis. She is started on broad spectrum antibiotics and taken for an urgent open cholecystectomy. The procedure went well, and it was without any immediate complications.<br><br>On post-operative day #1, you are paged by nursing after she was noted to have a brief run of asymptomatic atrial fibrillation picked up on telemetry. On detailed review of her telemetry | **Question 1**: Yes, additional monitoring, testing, and potentially treatment might be recommended given the episodes of atrial fibrillation (AF), even if brief and asymptomatic. Additional testing could include an echocardiogram to assess her cardiac structure and function, as AF can be associated with structural heart disease. Electrolyte monitoring and correction of any abnormalities, as well as assessing thyroid function, would also be prudent.<br><br>**Question 2a**: The decision to start therapeutic anticoagulation would be influenced by her risk of stroke, assessed by tools like the CHA2DS2-VASc score, and balanced against her bleeding risk (assessed by tools like HAS-BLED). Given the brief and asymptomatic nature of her AF episodes, and without further risk factor assessment, it's challenging to make a definitive recommendation without additional patient-specific information.<br><br>**Question 2b**: Factors influencing this decision include the duration and frequency of AF episodes, her stroke risk (based on age, presence |

| | |
|---|---|
| data, in the last 24 hours she has experienced 3 brief episodes of atrial fibrillation, each lasting 15-20 seconds.<br><br>From the perspective of her cholecystitis and peritonitis, she is clinically improving, has had no fevers or chills since the surgery, and she is tolerating a liquid diet. Her blood pressure and heart rate are currently normal. Her labs are notable only for a downtrending leukocytosis. Both a preoperative ECG and a current ECG show normal sinus rhythm without any significant abnormalities. She has no prior echocardiogram. She is currently on no anticoagulation or antiplatelet medications.<br><br>**QUESTION 1**: In addition to continuing telemetry monitoring, would you recommend any additional monitoring, testing, or treatment at this time? If so, what?<br><br>There are no episodes of atrial fibrillation on either post-operative days #2 or #3. However, on post-operative day #4, the day prior to anticipated discharge, the patient again develops another episode on telemetry that is confirmed by ECG. This episode is also completely asymptomatic. It lasts two hours, during which the ventricular rate ranges from 70 to 105 beats/min. Her blood pressure is not affected. It spontaneously converts back to normal sinus rhythm.<br><br>**QUESTION 2a**: Would you recommend to this patient that she be started on therapeutic anticoagulation?<br><br>**QUESTION 2b**: What factors would influence your decision?<br><br>**QUESTION 3**: If the decision was made to start the patient on therapeutic anticoagulation on hospital day #4 (i.e. immediately after the asymptomatic, 2 hour long episode of a-fib), what would be the best choice for anticoagulation?<br><br>**QUESTION 4a**: The patient is about to be discharged. Would you discharge her with an ambulatory ECG monitor? | of hypertension, diabetes, heart failure, or history of stroke/TIA), and her bleeding risk.<br><br>**Question 3**: If the decision was made to start anticoagulation, the best choice would depend on her renal function, risk of bleeding, and potential for drug-drug interactions. Given her stable renal function (CKD stage II), a direct oral anticoagulant (DOAC) like apixaban or dabigatran could be preferred for their ease of use and favorable risk profile compared to warfarin, assuming no contraindications.<br><br>**Question 4a**: Yes, discharging her with an ambulatory ECG monitor could be advisable to better characterize the frequency, duration, and potential triggers of her AF episodes.<br><br>**Question 4b**: A type of monitor chosen could be a Holter monitor for continuous recording over a few days or an event recorder for longer monitoring if episodes are infrequent.<br><br>**Question 4c**: Considerations include the frequency of her AF episodes, the patient's ability to use the device properly, and the clinical question being answered (e.g., burden of AF, symptom-rhythm correlation).<br><br>**Question 4d**: Information from the monitor would guide further management, such as the need for and type of anticoagulation, the potential benefit of antiarrhythmic drugs, and whether any lifestyle modifications or further interventions are warranted. |

| |
|---|
| **QUESTION 4b**: If so, what type of monitor would you choose?<br><br>**QUESTION 4c**: What factors should be considered in the choice of monitor for this patient?<br><br>**QUESTION 4d**: What would you do with the information from the monitor? |

**Supplement 5: Comparisons of the Primary and Secondary Outcomes by GPT Alone vs Physicians with GPT-4 and Physicians with Conventional Resources Only (Scores standardized to 0-100)**

| | | | | **Difference from GPT Alone** (based on Generalized Mixed Effect Model, 95% Confidence Interval, *p-value*) | |
|---|---|---|---|---|---|
| **Variable** | **Physicians+GPT-4**, N = 178[1] | **Physicians + Conventional Resources Only**, N = 197[1] | **GPT Alone**, N = 25[1] | **Physicians+GPT-4 vs GPT-4 Alone** | **Physicians + Conventional Resources Only vs GPT-4 Alone** |
| **Primary Outcome** | | | | | |
| **Total Score (n)** | 178 | 197 | 25 | -0.9 (-9.0 to 7.2), p=0.8 | -7.3 (-15.4 to 0.7), p=0.074 |
| Mean (SD) | 43.0 (17.3) | 35.7 (15.5) | 43.7 (14.2) | | |
| Median [IQR] | 41.3 [30.6 to 54.1] | 34.4 [22.5 to 47.8] | 41.8 [32.5 to 51.9] | | |
| **Secondary Outcomes** | | | | | |
| **Management (n)** | 178 | 197 | 25 | -2.7 (-10.4 to 5.1), p=0.5 | -8.7 (-16.5 to -1.0), p=0.028 |
| Mean (SD) | 40.5 (19.1) | 33.4 (17.3) | 42.7 (17.8) | | |
| Median [IQR] | 37.5 | 30.0 [19.3 to 45.5] | 41.8 | | |

| | | | | | |
|---|---|---|---|---|---|
| | [26.8 to 52.4] | | [29.5 to 55.0] | | |
| **Factual (n)** | 69 | 78 | 10 | 35.5 (10.1 to 61), p=0.007 | 26.3 (1.1 to 51.6), p=0.042 |
| Mean (SD) | 62.9 (37.6) | 53.8 (39.6) | 27.5 (31.1) | | |
| Median [IQR] | 75.0 [37.5 to 100.0] | 56.2 [15.6 to 100.0] | 18.8 [0.0 to 56.2] | | |
| **Diagnostic (n)** | 72 | 77 | 10 | 13.6 (-4.5 to 31.7), p=0.14 | 1.5 (-16.5 to 19.5), p=0.9 |
| Mean (SD) | 56.8 (37.6) | 45.8 (26.7) | 44.2 (33.3) | | |
| Median [IQR] | 66.7 [29.2 to 83.3] | 50.0 [33.3 to 66.7] | 41.7 [16.7 to 72.9] | | |
| **Specific (n)** | 178 | 197 | 25 | -4.4 (-12.5, 3.7), p=0.3 | -10.5 (-18.6 to -2.5), p=0.011 |
| Mean (SD) | 42.4 (20.2) | 34.9 (17.9) | 45.9 (21.6) | | |
| Median [IQR] | 42.6 [28.1 to 57.4] | 35.2 [20.8 to 48.5] | 51.9 [30.6 to 61.8] | | |

| | | | | | |
|---|---|---|---|---|---|
| **General (n)** | 70 | 80 | 10 | 5.3 (-4.2 to 14.8), p=0.3 | 2.0 (-7.5 to 11.4), p=0.7 |
| Mean (SD) | 29.4 (15.0) | 26.5 (13.0) | 24.8 (12.0) | | |
| Median [IQR] | 27.3 [18.2 to 39.8] | 24.6 [17.5 to 33.3] | 26.2 [12.5 to 34.2] | | |
| **Time Spent in Seconds (n)** | 178 | 197 | N/A | 696.2 (481.0 to 911.5), p<0.001 | 577.0 (362.3 to 791.7), p<0.001 |
| Mean (SD) | 801.5 (417.2) | 690.2 (372.4) | N/A | | |
| Median [IQR] | 719.8 [514.6 to 1,010.2] | 570.9 [452.9 to 814.9] | N/A | | |

**Supplement 6: Comparisons of the Primary Outcome by GPT Alone vs Physicians with GPT-4 and with Conventional Resources Only (Total score standardized to 0-100)**

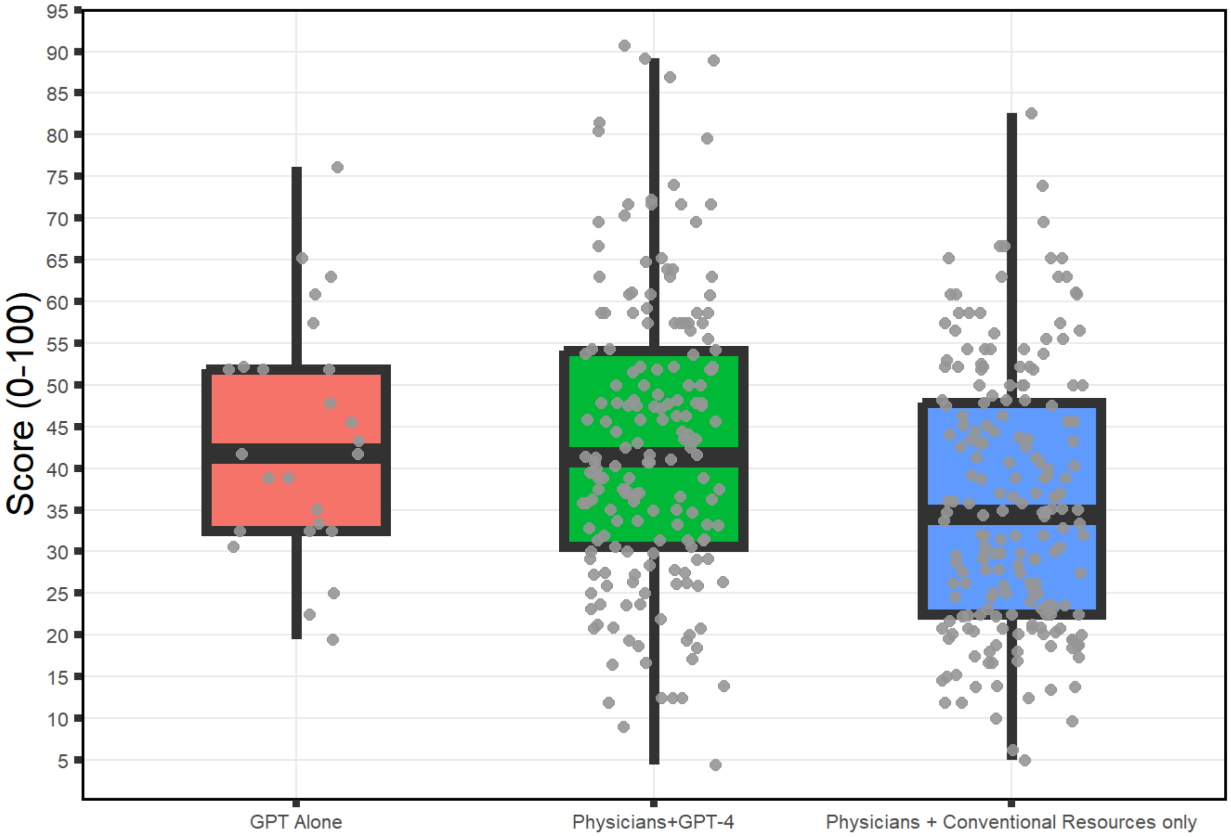

**Supplement 7: Comparisons of Time Spent by Physicians with GPT-4 and Physicians with Conventional Resources Only**

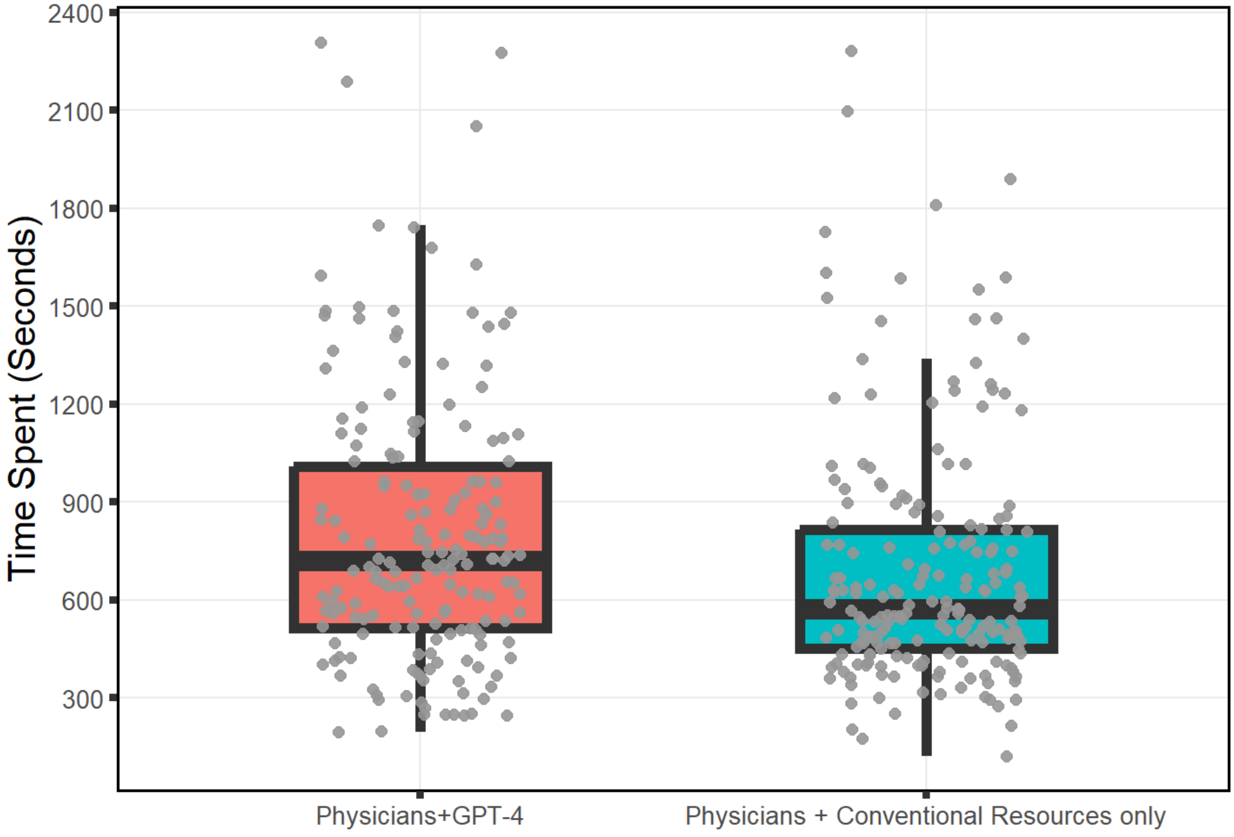
